# School bullying and social adaptation in Chinese adolescents: A multiple mediation model of self-disclosure and school connectedness

**DOI:** 10.1101/2022.06.10.22276238

**Authors:** Guo-Xing Xiang, Xiong Gan, Pin-Yi Wang, Rui-Jin Zhang, Xin Jin, Hao Li

## Abstract

**Background:** When it comes to the occurrence place of bullying behavior, school can never be ignored because adolescents spend a lot of time in school. School bullying has become a significant social issue among young generations, which influences their healthy growth. However, our understanding of the effects of school bullying is still limited. So, the present study aims to investigate how school bullying affects adolescent social adaptation.

**Methods:** A sample of 434 Chinese adolescents (56.9% females; *M*_*age*_=13.07 years, *SD*=0.93) participated in the survey. Structural equation modeling was adopted to assess the hypothesized model.

**Results:** The results indicated that school bullying had a direct effect on negative social adaptation rather than positive social adaptation. Moreover, self-disclosure and school connectedness mediated the relationship between school bullying and social adaptation, separately and sequentially.

**Conclusions:** The present study combines self-disclosure and school connectedness into a multiple mediation model, highlighting the importance of individual and environmental factors in the effects of school bullying on adolescents’ social adaptation. Practically, the current findings may provide some guidance for the prevention and intervention of school bullying and the promotion of social adaptation in adolescents.

## 1. Introduction

Bullying is defined as aggressive behavior that happens intentionally and repeatedly, which usually involves an imbalance of power [1]. Bullying can take different forms, including physical bullying (e.g., pushing), verbal bullying (e.g., teasing), social exclusion (e.g., ignoring someone) and other indirect forms (e.g., spreading rumors), and it always includes at least one bully and one victim [2]. An increasing number of studies have demonstrated that bullying can result in developmental problems and other negative outcomes, including mental disorders (e.g., anxiety and depression), personality disorders (e.g., antisocial personality), behavioral problems (e.g., crime), poor health, poor wealth, and lower subjective well-being [3-8]. Therefore, understanding how bullying affects adolescents’development is crucial in the prevention and intervention of bullying.

When it comes to the occurrence place of bullying behavior, school can never be ignored because adolescents spend a lot of time in school. A recent survey has reported that one in three (32%) students experience bullying by peers at school [9]. Therefore, more and more scholars pay attention to school bullying, where they have made great achievements in its antecedents (e.g., neuroticism, loneliness, and parental behavioral control) [10, 11] and consequences (e.g., substance use) [12]. Few studies, however, focus on the relationship between school bullying and social adaptation, which is an important indicator of individual socialization. To address this research gap, the present study aims to investigate the relationship between school bullying and social adaptation and explore the underlying mechanisms between them. This will contribute to adolescents’ social adaptation by preventing them from suffering school bullying.

### 1.1 School Bullying and Social Adaptation

Social adaptation refers to the dynamic process by which individuals change attitudes and behaviors in order to maintain a harmonious and balanced relationship with their social environment [13]. According to the area-function conceptual model of adolescent social adaptation, social adaptation consists of two functional states: positive and negative social adaptation, and good social adaptation is comprehensively shaped by two aspects: a high level of positive social adaptation and a low level of negative social adaptation [14].

Positive social adaptation involves behaviors promoting healthy development and meeting social norms, such as self-affirmation, active coping, and prosocial tendency [14]. Empirical evidence has indirectly indicated the potential association between school bullying and positive social adaptation. For instance, self-esteem and self-efficacy (both included in self-affirmation) are negatively correlated with school bullying [15-18]. Similarly, a negative association between prosocial behaviors and school bullying has also been reported among New Zealand students [19]. Moreover, recent studies have revealed that active coping can buffer the detrimental influence of bullying [20, 21]. In sum, this literature jointly suggests that adolescents with experience of school bullying tend to develop lower levels of self-affirmation, active coping, and prosocial tendencies. Given that the three facets of positive social adaptation are negatively associated with school bullying, it is reasonable to assume that school bullying will negatively predict positive social adaptation.

Negative social adaptation, on the other hand, refers to behaviors unconducive to individual growth, such as self-trouble, social alienation, and social withdrawal [14]. Prior studies have revealed that social alienation can be positively predicted by school bullying [22, 23]. Moreover, compared with non-school bully/victim students, adolescents involved in bullying have a higher level of social withdrawal [24]. This indirect evidence, to a certain degree, implies a positive association between school bullying and negative social adaptation. In addition, developmental contextualism emphasizes that people have an impact on the environment and that individual development will be shaped by the environment as well [25]. Bullying perpetration can be regarded as a kind of negative influence of adolescents on their peers in the same environment, which will trigger a series of adverse effects on the social adaptation of bullies. For instance, bullies will be punished by their school and family, and their peers will not be willing to develop friendships with them. In the long term, individuals who are involved in bullying will develop more maladaptive behaviors such as theft, violence, binge-drinking, and smoking [26, 27]. Accordingly, it is reasonable to assume that school bullying will positively predict negative social adaptation. Overall, involvement in bullying perpetration at school will be destructive to adolescents’ adaptive development. Thus, the present study hypothesizes that school bullying will negatively predict positive social adaptation and positively predict negative social adaptation (H1).

### 1.2. Mediating Effect of Self-Disclosure

It is worthwhile to note that the relationship between school bullying and social adaptation may be direct or indirect. The present study attempts to further explore the potential mediators between the aforesaid relationship. Self-disclosure, a possible mediator, is defined as the behavior of revealing private information about oneself to others [28]. Studies on self-disclosure usually focus on its breadth and depth. The former means the wide range of content disclosed, while the latter represents the degree of privacy of information disclosed [29]. Self-disclosure plays an important role in the development of relationships with family, friends, and partners [30]. Likewise, self-disclosure might also have an essential impact on the association between school bullying and social adaptation, which will be discussed in the following two paragraphs.

Abundant studies have confirmed that self-disclosure is closely linked to various forms of bullying. For instance, Georgiou and Stavrinides [31] found that early adolescents with bullying experience at school show less self-disclosure. Similarly, among middle school girls, Jones et al. [32] reported that girls involved in school bullying are less likely to disclose themselves. Both the findings support the negative effect of school bullying on self-disclosure. In addition, an analogous negative relationship is also revealed between cyberbullying and self-disclosure [33-36]. Considering the prior evidence, it is reasonable to propose that school bullying will negatively predict self-disclosure among adolescents.

Besides, self-disclosure is also tightly associated with social adaptation throughout a lifetime. According to social penetration theory, self-disclosure is of vital importance in the development of interpersonal relationships since the process of self-disclosure will strengthen the relationship in a penetrating way [37], which in turn avoids maladaptive outcomes such as social alienation and withdrawal. Empirical studies have also demonstrated a significantly positive association between self-disclosure and social adaptation among adults and university students [38-40]. Moreover, Yu[41] revealed a similar finding that adolescents with a high level of self-disclosure are more likely to develop better social adaptation. Therefore, based on the above theory and evidence, it is appropriate to deduce that self-disclosure will significantly predict more positively adaptive outcomes and fewer maladaptive outcomes.

Given the independent associations between self-adaptation and the two variables, the present study attempts to further assess the mediating effect of self-disclosure on the relationship between school bullying and social adaptation. In a study of overweight adolescents, Adams and Cantin [42] revealed the buffering effect of self-disclosure on the negative impacts of peer bullying on depression, which supports the assumption of the mediating effect of self-disclosure in the present study. Consequently, the present study hypothesizes that self-disclosure will significantly mediate the relationship between school bullying and social adaptation (H2).

### 1.3. Mediating Effect of School Connectedness

School connectedness, in addition to self-disclosure, might also play a mediating role in the relationship between school bullying and social adaptation. School connectedness refers to the emotional connection established between individuals and schools, as well as the people in school, which reflects the students’ sense of belonging and identity to school, and their feelings of being cared for, recognized, and supported in school [43]. School connectedness is an effective factor in the prevention of developmental problems [44]. Therefore, it is worthwhile to evaluate its protective functions in the present topic.

Prior literature has provided both indirect and direct evidence linking school connectedness with the two major variables. In terms of the association between school bullying and school connectedness, Skues et al. [45] found that, with higher engagement in school bullying, adolescents will feel unsafe and unfriendly, and tend to disconnect with their peers, teachers, and schools. Moreover, Diaz et al. [46] and O’Brennan and Furlong [47] repeatedly revealed the stably negative association between school bullying and connectedness. Therefore, it is reasonable to believe the negative influence of school bullying on school connectedness among adolescents. As for the association between school connectedness and social adaptation, a longitudinal study suggests that school connection can predict a lower level of adaptation problems in early adolescents one year later [48]. Additionally, Yin et al. [49] indicated that middle school students who are highly connected with their schools are more easily adapted to society. School is a micro-society with some characteristics of real society, and it provides a platform for adolescents to rehearse and prepare. So, the connection with school reflects the social function of students to a certain extent. It is not too difficult for adolescents with strong social functions to adapt to today’s complex society. Overall, school bullying could negatively influence school connection, which in turn would predict less positive social adaptation and more maladaptive outcomes.

Moreover, the risk-buffering model highlights that protective factors can alleviate the negative influence of risk factors on individual growth [50]. Liu et al. [51] revealed that school connectedness can buffer the adverse effect of bullying on psychological well-being. Furthermore, Loukas and Pasch [52] indicated that school connectedness alleviates the impact of bullying victimization on subsequent adjustment problems of girls in early adolescence. Both of the empirical findings point to the protective function of school connectedness between risk factors and individual development. Therefore, the present study hypothesizes that school connectedness will mediate the relationship between school bullying and social adaptation (H3).

### 1.4. Self-Disclosure and School Connectedness

As reviewed above, both self-disclosure and school connectedness are associated with school bullying and social adaptation. Furthermore, there might be another potential association between self-disclosure and school connectedness. The disclosure reciprocity effect suggests that, in social interaction, one’s self-disclosure will trigger the other’s self-disclosure [53]. This process is necessary in the development of a relationship because knowing each other is a prerequisite for establishing a relationship. In school, students disclose themselves to let peers and teachers understand them, allowing them to feel cared for, recognized, and supported. Long term, students may discover similarities among themselves, resulting in a sense of belonging and identity to the group and school. That is, they are connected to their schools.

Besides, social penetration theory suggests that telling others about one’s private information is beneficial to establishing a good connection [37]. Empirical research has found that disclosing will make the interaction or relationship more intimate and stable [54, 55]. For instance, Lee et al. [56] revealed that self-disclosure can improve received social support, which increases an individual’s feeling of well-being. Similarly, Zhang’s study [57] also suggested that disclosure is positively related to social support. So, self-disclosure makes adolescents feel supported, which is helpful in building connections with peers and teachers. Adolescents’ self-disclosure in social interaction can positively influence the content of connectedness in school.

Overall, school bullying could negatively influence adolescents’ self-disclosure and school connectedness, and all of them have effects on adolescents’ social adaptation. Moreover, self-disclosure could foster a better level of connection with school. In light of these complicated associations, the present study attempts to further explore whether self-disclosure and school connectedness will function as a chain mediating effect in the aforesaid relationship. That is, engagement in school bullying will make adolescents feel unsafe, and they will not be willing to share private information, so it is difficult for them to build intimate connections with their peers, teachers, and schools, which in turn is detrimental to their adaptative development. Accordingly, the present study hypothesizes that self-disclosure and school connectedness may have a serial mediating effect on the relationship between school bullying and social adaptation (H4).

### 1.5. The present study

With the combination of several theories and previous evidence, we realized the complex relationship between school bullying and social adaptation among adolescents. Little research, however, has compared the direct and indirect associations in the aforesaid relationship, as well as the comparation of three possibly mediating pathways. This comparison, in practice, will be in the service of better assisting adolescents to circumvent the adverse impacts of school bullying on their adaptative development. Therefore, the present study aims to fill this knowledge gap and construct a serial mediation model with hypotheses that (Figure 1):

**Figure 1.**
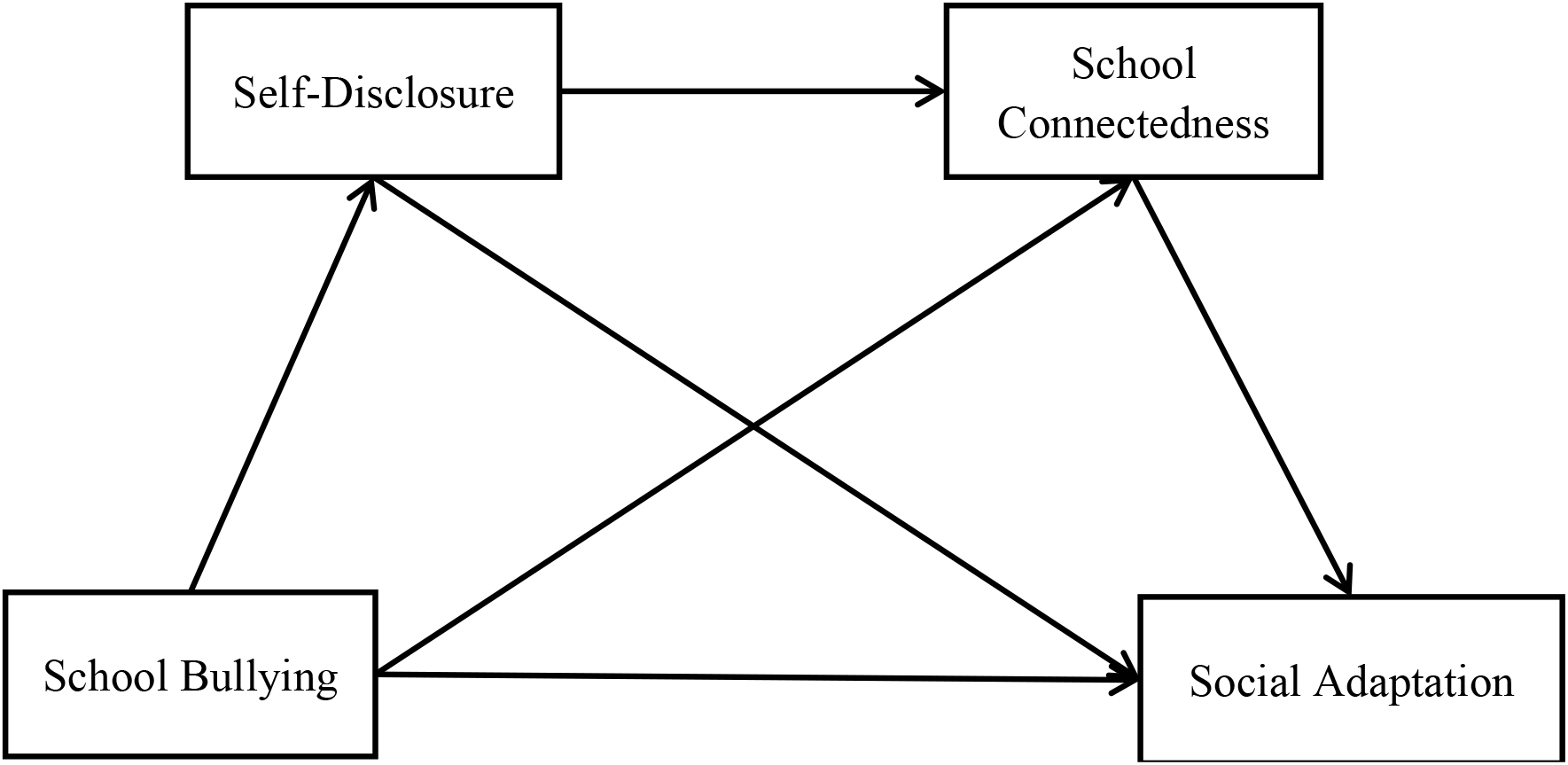
The relationship between school bullying and social adaptation among adolescents.

***H1:*** *School bullying will negatively predict positive social adaptation and positively predict negative social adaptation*.

***H2:*** *Self-disclosure will mediate the relationship between school bullying and social adaptation*.

***H3:*** *School connectedness will mediate the relationship between school bullying and social adaptation*.

***H4:*** *Self-disclosure and school connectedness may have a serial mediating effect on the relationship between school bullying and social adaptation*.

## 2. Method

### 2.1. Participants and Procedure

A total of 434 junior high school students were recruited from Guizhou province, southwest China. Of these, 247 (56.9%) were female. Their ages ranged from 11 to 17 years old, and the average age was 13.07 years (*SD*=0.93). This study was approved by the Research Ethics Committee of the College of Education and Sports Sciences, Yangtze University. After obtaining informed consent verbally, students were asked to finish a questionnaire on school bullying, social adaptation, school connectedness, self-disclosure, and several basic information such as gender and age within half an hour. At the end, researchers collected all the questionnaires and thanked them for participating. Students were informed that participation was entirely voluntary, and they did not receive any compensation for participation.

### 2.2. Measures

#### School Bullying

Adolescent school bullying was measured by the Bully Subscale taken from the Adapted Chinese Version of the Revised Olweus Bully/Victim Questionnaire (ROBVQ) [58, 59]. It consists of seven items evaluated on a four-point scale (0 = never; 4 = several times a week), assessing four kinds of bully behaviors, including physical bullying, verbal bullying, social bullying, and cyberbullying. One example item is, “I posted some students’ secrets, private or embarrassing photos on the Internet.” A higher score indicates more bullying behaviors. In this study, its Cronbach’s alpha coefficient was 0.833.

#### Social Adaptation

Adolescent social adaptation was measured by the Adolescent Social Adjustment Assessment Scale (ASAAS) [14]. This scale has 50 items in total, eight dimensions including self-affirmation, active coping, prosocial tendency, acting efficiency, self-trouble, social alienation, violations, and social withdrawal. The first four dimensions are considered the Subscale of Positive Social Adaptation, with a higher score indicating better positive social adaptation; the other four dimensions are considered the Subscale of Negative Social Adaptation, with a higher score meaning worse negative social adaptation. One example item is “I am not happy at all”. Responses range from “1 = totally inconsistent” to “5 = totally consistent”. Its Cronbach’s alpha coefficient was 0.703 in this study.

#### Self-disclosure

Adolescent self-disclosure was measured by the Adolescent Self-Disclosure with Peers Questionnaire (ASDPQ) [60]. This questionnaire includes 37 items, seven dimensions such as interests and study experience. One example item is “I will tell my classmates who are not so close to me about my exam results”. Items were evaluated on a five-point scale (1 = almost nothing; 5 = anything), with a higher score indicating a better level of self-disclosure. Its Cronbach’s alpha coefficient was 0.977 in this study.

#### School Connectedness

Adolescent school connectedness was measured by the Adolescent School Connectedness Scale (ASCS) [61]. It contains 10 items and three dimensions, such as teacher support, peer support, and school belonging. One example is, “I feel happy and safe in my school.” Items were rated on a five-point scale, ranging from “1 = totally inconsistent” to “5 = totally consistent”. A higher score indicates a better connection with school. In this study, its Cronbach’s alpha coefficient was 0.832.

### 2.3. Data Analysis

SPSS 26.0 and PROCESS macro were used for data analysis [62]. First, preliminary analyses such as descriptive and correlational statistics were performed in SPSS 26.0 to assess whether the variables were associated with each other. Then, mediation analyses were conducted through model six in PROCESS macro to examine the mediating roles of school connectedness and self-disclosure between school bullying and social adaptation.

## 3. Results

### 3.1. Common Method Biases Analyses

Since the data were from participants’ self-report questionnaire, it is necessary to avoid common method biases. And the Harman single factor method was recommended to detect it [63]. Results showed that there were 39 factors with a characteristic value greater than one. Additionally, the first factor explained a variation of 18.44%, less than the 40% critical value. That is, the results of the present study were less influenced by common method biases.

### 3.2. Descriptive Statistics and Correlation Analyses

Means and standard deviations for all variables are displayed in Table 1. Chi-square tests reported significant gender differences in school bullying (*χ*^*2*^ = 6.771, *p*<0.01). Boys had more bullying behaviors in school than girls. And *t* tests found that girls’ positive social adaptation (*t* = -2.037, *p*<0.05), self-disclosure (*t* = -3.109, *p*<0.01) and school connectedness (*t* = -3.893, *p*<0.001) were much better than boys’, while boys were the same as girls in negative social adaptation (*t* = 1.948, *p*> 0.05). Besides, grade differences were also significant in both positive (*t* = 2.429, *p*< 0.05) and negative (*t* = -2.693, *p*<0.01) social adaptation. Compared with eighth grade students, adolescents in seventh grade had a higher level of positive social adaptation and a lower level of negative social adaptation. Hence, gender and grade were considered as control variables in the following analyses.

**Table 1.** Descriptive statistics for key variables.

| Variable | Boys |  | Girls |  |
| --- | --- | --- | --- | --- |
|  | *M* | *SD* | *M* | *SD* |
| 1. SB | 0.134 | 0.341 | 0.061 | 0.239 |
| 2. PSA | 3.484 | 0.748 | 3.623 | 0.668 |
| 3. NSA | 2.169 | 0.698 | 2.045 | 0.622 |
| 4. SD | 2.414 | 0.702 | 2.622 | 0.671 |
| 5. SC | 3.872 | 0.798 | 4.134 | 0.606 |

**Note.** SB=School bullying, PSA= Positive social adaptation, NSA= Negative social adaptation, SD= Self-disclosure, SC= School connectedness.

Pearson correlation analysis was used to explore the association of key variables. As presented in Table 2, school bullying was negatively associated with positive social adaptation (*r* = -0.279, *p*<0.01), self-disclosure (*r* = -0.211, *p*<0.01), school connectedness (*r* = -0.399, *p*<0.01), while it was positively associated with negative social adaptation (*r* = 0.288, *p*<0.01). Positive social adaptation was positively correlated with self-disclosure (*r* = 0.252, *p*<0.01) and school connectedness (*r* = 0.592, *p*<0.01), but negatively correlated with negative social adaptation (*r* = -0.357, *p*<0.01). In addition, negative social adaptation had significantly negative relationships with self-disclosure (*r* = -0.161, *p*<0.01) and school connectedness (*r* = -0.435, *p*<0.01). A significantly positive association was found between self-disclosure and school connectedness (*r* = 0.239, *p*<0.01).

**Table 2.** Correlation analysis of key variables.

| Variable | 1 | 2 | 3 | 4 | 5 |
| --- | --- | --- | --- | --- | --- |
| 1. SB | 1 |  |  |  |  |
| 2. PSA | -0.279** | 1 |  |  |  |
| 3. NSA | 0.288** | -0.357** | 1 |  |  |
| 4. SD | -0.211** | 0.252** | -0.161** | 1 |  |
| 5. SC | -0.399** | 0.592** | -0.435** | 0.239** | 1 |

**Note.** SB=School bullying, PSA= Positive social adaptation, NSA= Negative social adaptation, SD= Self-disclosure, SC= School connectedness. \*\* $p < 0.01$ .

### 3.3. Mediation Effect Analyses

#### 3.3.1. Mediating Effects between School Bullying and Positive Social Adaptation

After data standardization, a serial mediation model was constructed to examine the possible impacts of self-disclosure and school connectedness on the relationship between school bullying and positive social adaptation. Results of regression analyses (Table 3) showed that school bullying could negatively predict positive social adaptation (*β* = -0.261, *t* = -5.618, *p*<0.001) and self-disclosure (*β* = -0.195, *t* = -4.115, *p*<0.001). Besides, school connectedness could be predicted by both self-disclosure (*β* = 0.146, *t* = 3.286, *p*<0. 01) and school bullying (*β* = -0.348, *t* = -7.819, *p*<0. 001). But when adding all the variables into the model, school bullying (*β* = -0.032, *t* = -0.745, *P*>0.05), rather than self-disclosure (*β* = 0.115, *t* = -2.871, *p*<0.01) or school connectedness (*β* = 0.550, *t* = 12.826, *p*<0. 001), failed to predict positive social adaptation. confidence interval, LL= lower limit, UL= upper limit.

**Table 3.**
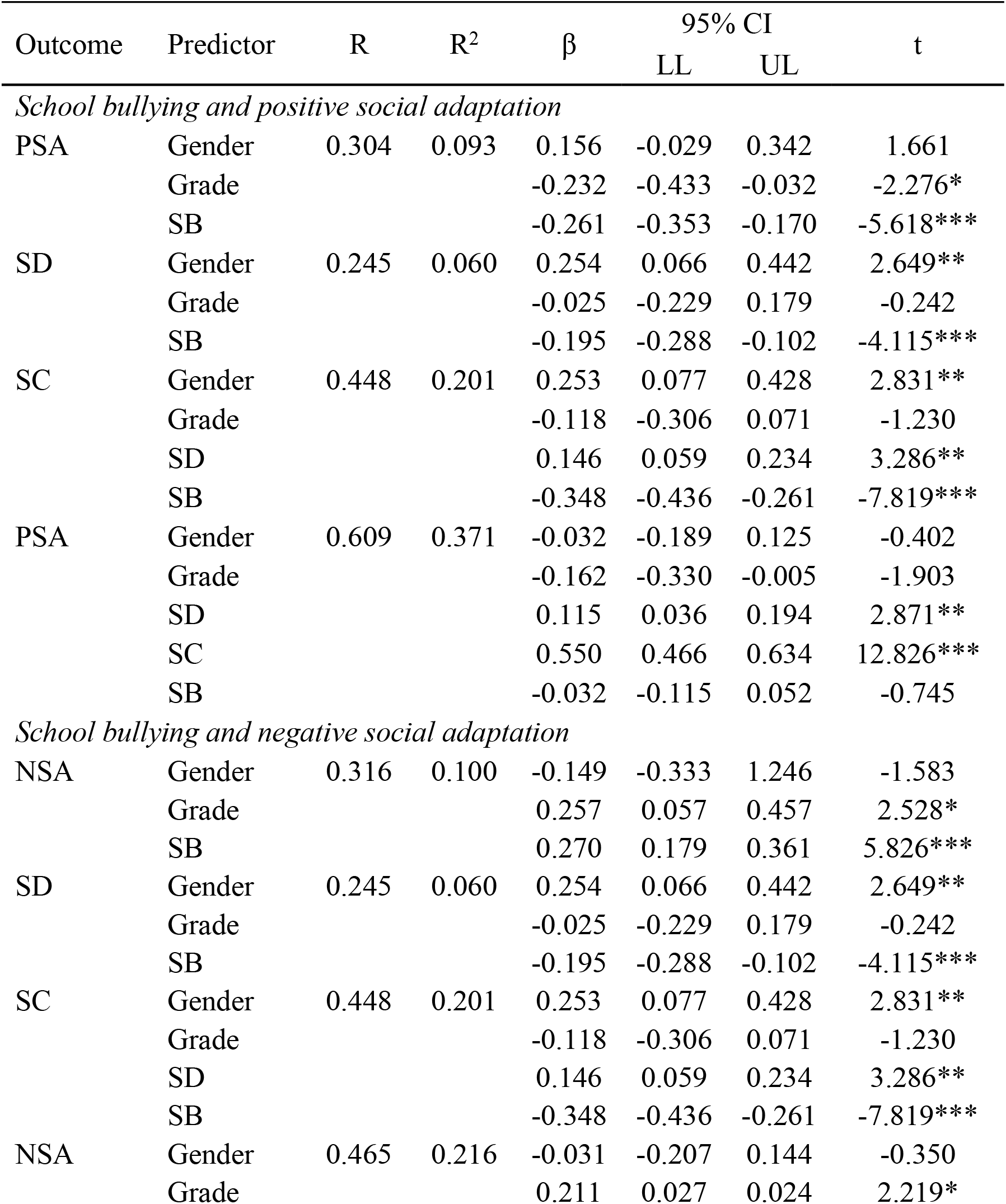
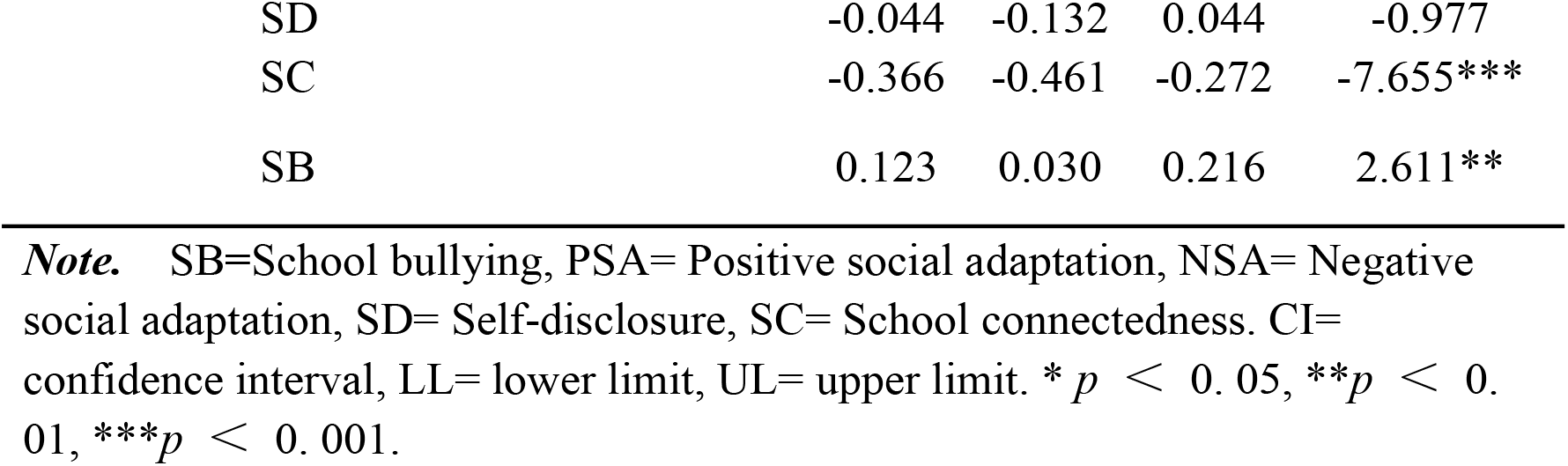
Regression analysis of pathways between school bullying and social adaptation.

Further analyses suggested that self-disclosure and school connectedness played mediating roles in the association between school bullying and positive social adaptation (Table 4). Figure 2 displays the direct and indirect pathways. Self-disclosure mediated the pathway from school bullying to positive social adaptation (*β* = -0.022, *95% CI* = [-0.044, -0.006]). In particular, school bullying could predict less self-disclosure, which further led to less positive social adaptation. Similarly, school bullying could also result in less positive social adaptation via decreasing school connectedness (*β* = -0.192, *95% CI* = [-0.257, -0.133]). In terms of the serial mediation effects, results suggested that school bullying reduced self-disclosure, which caused less school connection, when then further decreased positive social adaptation (*β* = -0.016, *95% CI* = [-0.029, -0.006]).

**Table 4.**
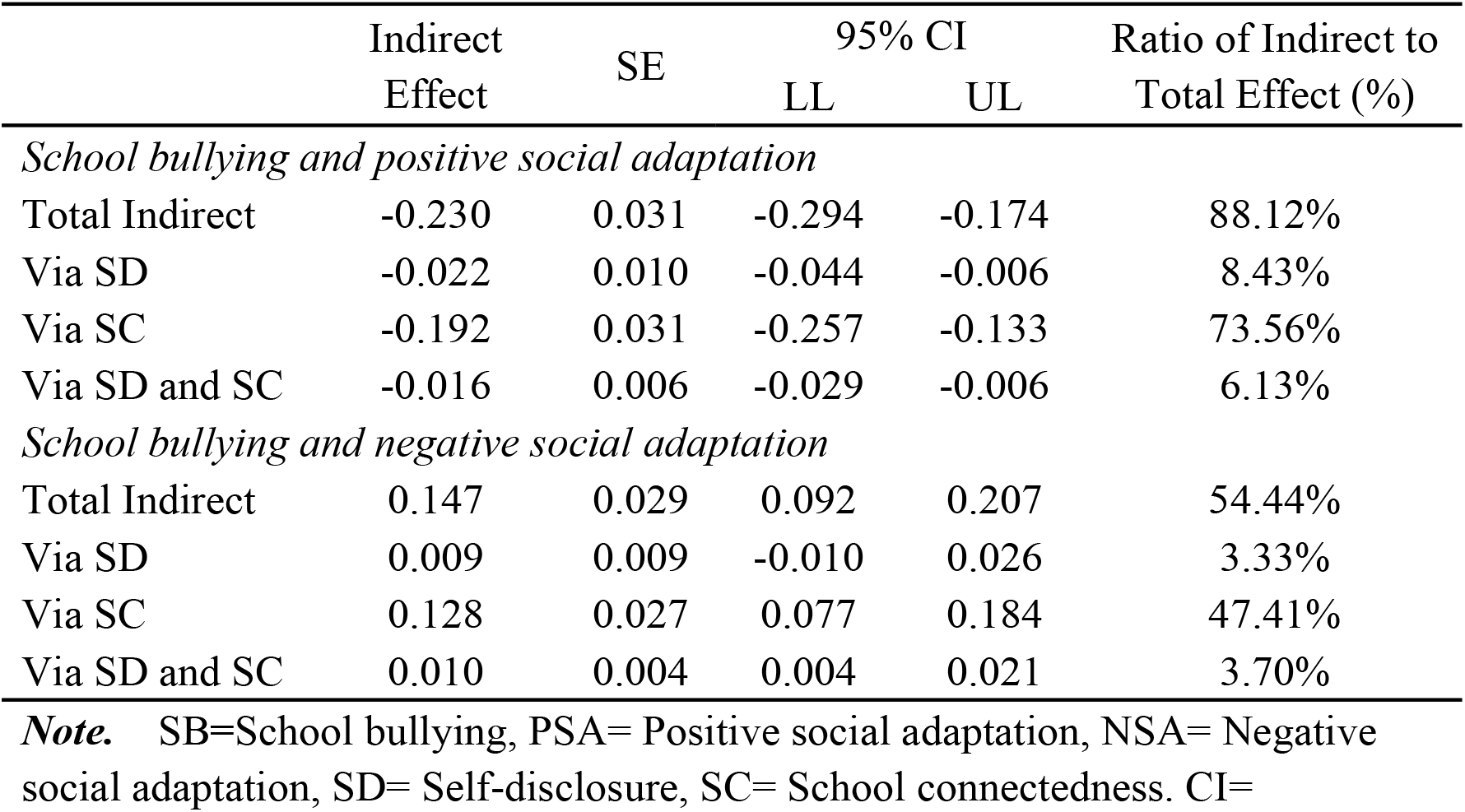
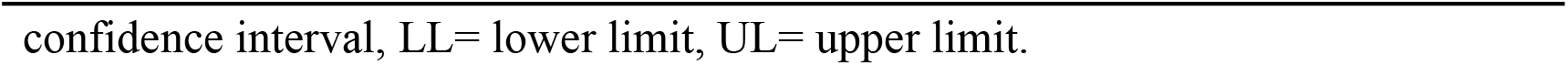
Standardized indirect effects from school bullying and social adaptation

|  | Indirect Effect | SE | 95% CI |  | Ratio of Indirect to Total Effect (%) |
| --- | --- | --- | --- | --- | --- |
|  |  |  | LL | UL |  |
| *School bullying and positive social adaptation* |  |  |  |  |  |
| Total Indirect | -0.230 | 0.031 | -0.294 | -0.174 | 88.12% |
| Via SD | -0.022 | 0.010 | -0.044 | -0.006 | 8.43% |
| Via SC | -0.192 | 0.031 | -0.257 | -0.133 | 73.56% |
| Via SD and SC | -0.016 | 0.006 | -0.029 | -0.006 | 6.13% |
| *School bullying and negative social adaptation* |  |  |  |  |  |
| Total Indirect | 0.147 | 0.029 | 0.092 | 0.207 | 54.44% |
| Via SD | 0.009 | 0.009 | -0.010 | 0.026 | 3.33% |
| Via SC | 0.128 | 0.027 | 0.077 | 0.184 | 47.41% |
| Via SD and SC | 0.010 | 0.004 | 0.004 | 0.021 | 3.70% |

**Note.** SB=School bullying, PSA= Positive social adaptation, NSA= Negative social adaptation, SD= Self-disclosure, SC= School connectedness. CI=
confidence interval, LL= lower limit, UL= upper limit.

**Figure 2.**
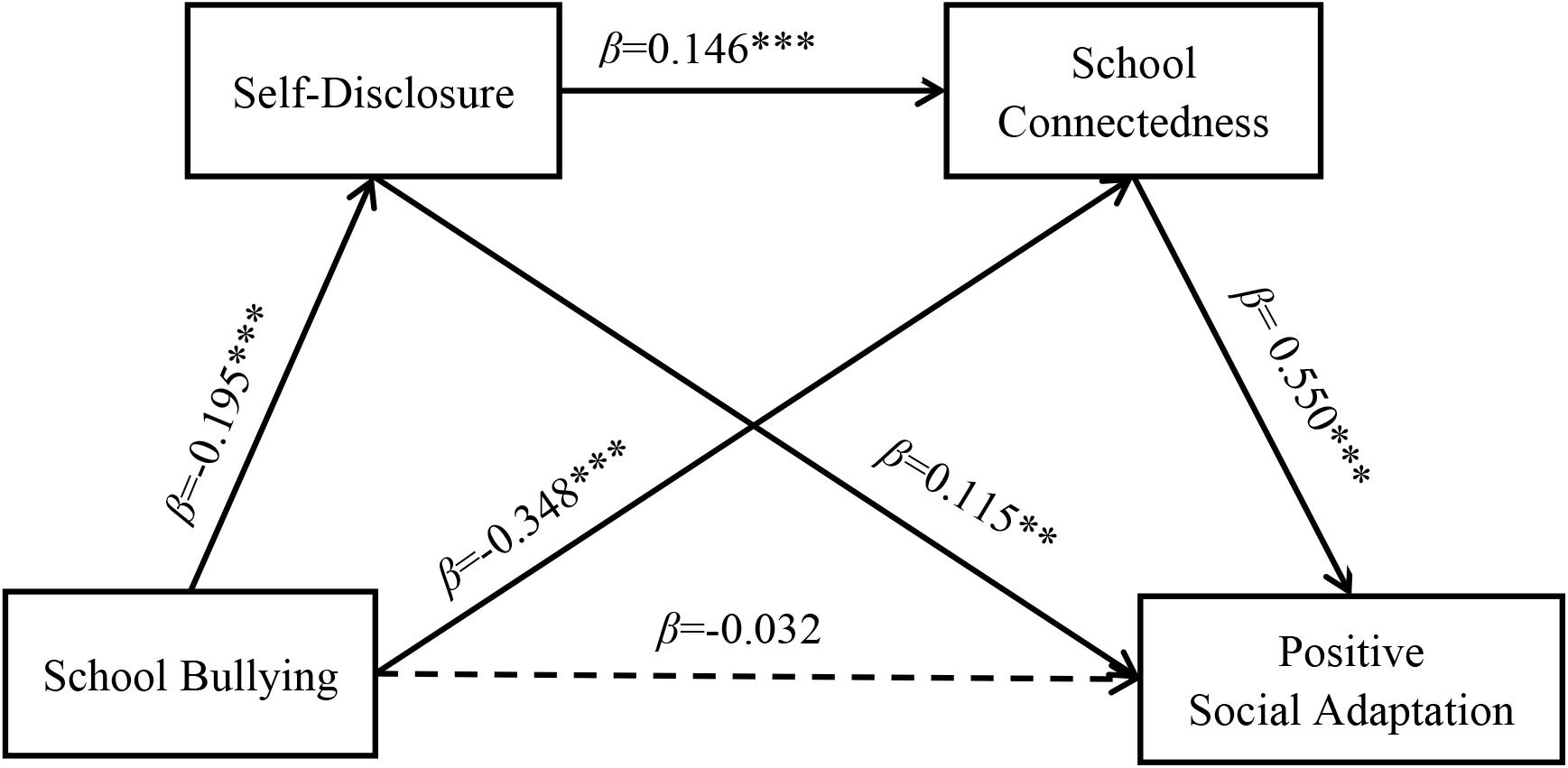
The serial mediation model between school bullying and positive social adaptation Note. \*\**p* < 0. 01, \*\*\**p* < 0.001.

#### 3.3.2. Mediating Effects between School Bullying and Negative Social Adaptation

Likewise, another serial mediation model was also constructed to test the possible influence of self-disclosure and school connectedness on the association between school bullying and negative social adaptation. Results of regression analyses (Table 3) revealed that school bullying could predict negative social adaptation (*β* = 0.270, *t* = 5.826, *p*<0.001), self-disclosure (*β* = -0.195, *t* = -4.115, *p*<0.001) and school connectedness (*β* = -0.348, *t* = -7.819, *p*<0. 001). Self-disclosure had a significantly positive impact on school connectedness (*β* = 0.146, *t* = 3.286, *p*<0. 01). When taking all those variables into the model, school connectedness (*β* = -0.366, *t* = -7.655, *p*<0. 001) and school bullying (*β* = 0.123, *t* = 2.611, *p*<0.01) could predict negative social adaptation significantly.

Further pathway analyses reported that self-disclosure and school connectedness played mediating roles in the association between school bullying and negative social adaptation (Table 4). As displayed in Figure 3, school bullying had an effect on negative social adaptation via school connectedness (*β* = 0.128, *95% CI* = [0.077, 0.184]). Moreover, a chain mediating path from self-disclosure to school connectedness was significant between school bullying and negative social adaptation (*β* = 0.010, 9*5% CI* = [0.004, 0.021]).

**Figure 3.**
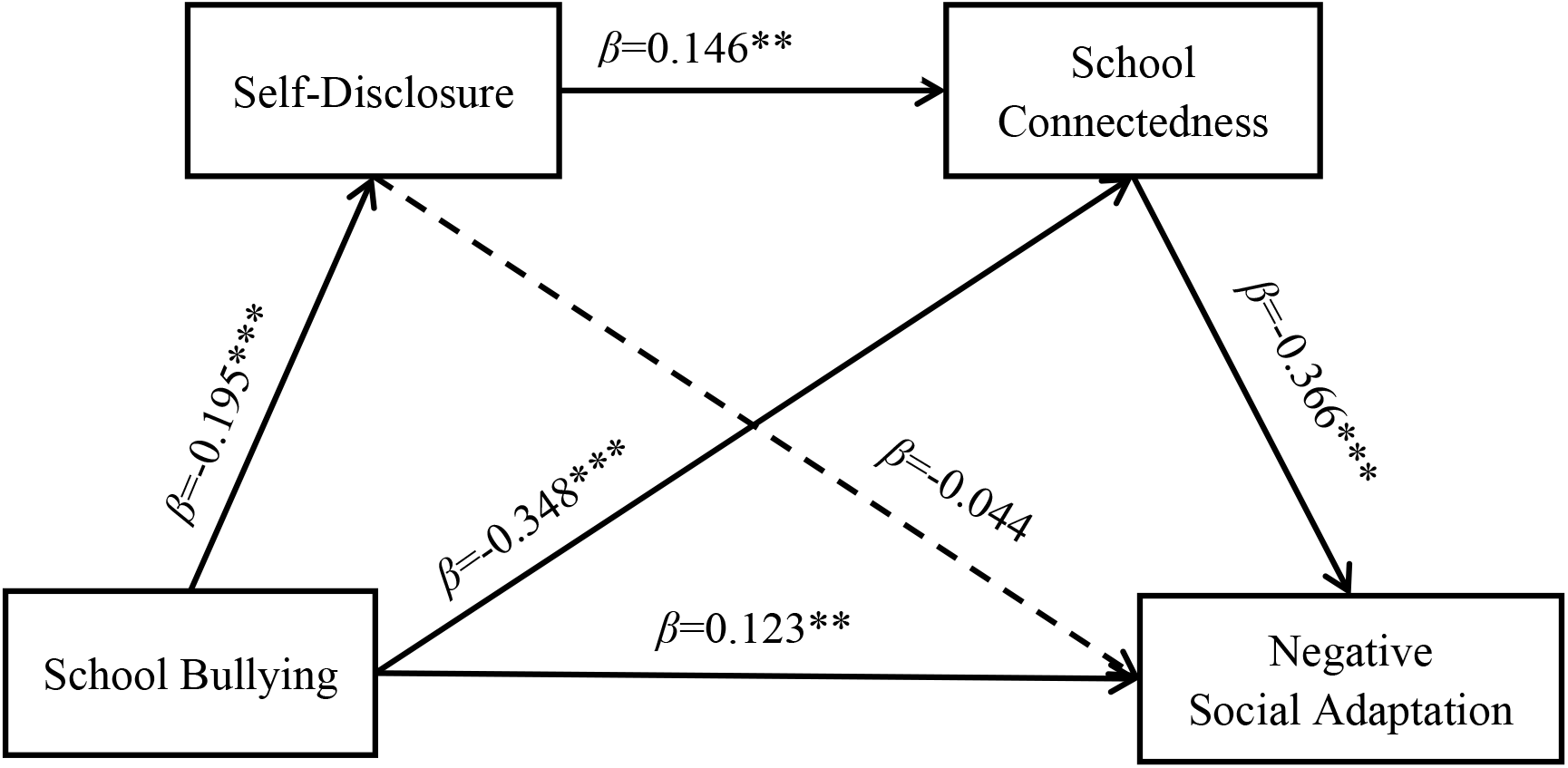
The serial mediation model between school bullying and negative social adaptation Note. **p<0. 01, ***p<0.001.

## 4. Discussion

The present study attempts to figure out the complex relationship between school bullying and social adaptation during adolescence by simultaneously assessing the mediation effects of self-disclosure and school connectedness in a serial mediation model. Structural equation modeling was performed to analyze data from a sample of Chinese students, and the findings generally supported the hypothesized model. School bullying had an influence on social adaptation through the mediating pathways of self-disclosure and school connectedness. Below, these findings will be discussed in more detail.

To start, the first hypothesis, that there is a significant connection between school bullying and social adaptation, was supported among Chinese adolescents. Specifically, school bullying could negatively predict positive social adaptation while it could positively predict negative social adaptation. That is, compared with adolescents without experiencing school bullying, those who are involved in bullying others are more likely to develop less positive social adaptation outcomes and more negative social adaptation outcomes. This finding is consistent with previous research, which indicates a significant relationship between school bullying and specific content of social adaptation (e.g., self-affirmation, prosocial behavior, and social alienation) [16, 19, 23]. Moreover, this finding extends existing literature by providing direct evidence for the relationship between school bullying and adolescents’ social adaptation.

One possible explanation of this finding given by developmental contextualism is that individuals have an impact on their environment. Meanwhile, individual development will also be affected by the environment [25]. Adolescent bullying will have negative impacts on victims, such as physical injury and psychological harm. In the meantime, bullying will have adverse effects on bullies as well. The most immediate consequence is that they will be punished, excluded, and lose friendships because their peers will feel threatened by them and keep a wide berth away from them. Thus, it will lead to negative social adaptation for bullies, such as social alienation. More seriously, bullies will develop the cognitive bias of believing that bullying others is a symbol of power, while actually bullying is considered a deviant behavior in violation of the moral rules. With this cognitive bias, it will be difficult for bullies to develop positive social adaptations such as prosocial behaviors.

Similar to Adams and Cantin’s [42] study of the buffering effect of self-disclosure, the present study revealed that self-disclosure mediated the relationship between school bullying and positive social adaptation, rather than negative social adaptation. That is, the second hypothesis partly received support. Adolescents involved in school bullying are less likely to disclose themselves, which is not beneficial to positive social adaptation. Theoretically, this finding integrates previous independent studies on the association between self-disclosure and school bullying and social adaptation [31, 32], by revealing its mediating effect. Besides, this finding provides an important pathway for practitioners to prevent adolescents from being influenced by school bullying.

According to the social penetration theory, self-disclosure is critical in the development of interpersonal connections and social adaptation [37]. Because self-disclose can contribute to information exchange, making oneself understood by others, and then others will respond to the information. The feedback, especially the positive one, can assist one in keeping or correcting behaviors. In the long term, adolescents will be more likely to ask for advice or feedback when they are facing difficulties. That is, they will cope in positive ways and take action efficiently. Moreover, they will gradually develop self-affirmation and confidence in themselves. All of these positive effects could promote positive social adaptation later. However, being involved in bullying others will make adolescents feel unsafe and unsupported, so they are not willing to disclose themselves. Thus, it is not possible for bullies to benefit from the feedback of self-disclosure. Their violent behaviors, as well as their cognitive biases, will not be corrected. As a result, bullies will face more maladaptive developmental problems. Additionally, although self-disclosure was negatively related to negative social adaptation and positively related to school bullying, it could not mediate the association between them. One possible reason is the masking effect of other pathways. Compared with the direct association and the mediating pathway through school connectedness, the mediating function of self-disclosure is too marginal to be significant. Even if it is remarkable in the association between school bullying and positive social adaptation, self-disclosure still plays a much smaller role compared to school connectedness.

In addition, school connectedness is another mediator between school bullying and social adaptation, which is in favor of the third hypothesis. Experiencing bullying others at school will make adolescents disconnect with their schools, which will lead to less positive social adaptation and more negative social adaptation. This finding is similar to the prior results of Liu et al.’s study [51] and Loukas and Pasch’s study [52]. Moreover, it is consistent with prior studies that school connectedness is closely associated with adolescent school bullying and social adaptation outcomes [48, 49, 51]. This finding extends previous independent studies on the association between self-disclosure and school bullying and social adaptation by revealing its mediating role. Besides, it also provides another pathway for practitioners to prevent adolescents from suffering the harmful consequences of school bullying.

Furthermore, a combination of social identity theory and Maslow’s hierarchy of needs theory could provide a better understanding of this finding [64, 65]. The membership of social groups is an important part of self-concept. In social interaction, people always strive to maintain a positive social identity to enhance their sense of self-worth. Additionally, individuals have a need for belonging and love. Therefore, in order to seek membership, a sense of belonging, and love, adolescents will spare no effort to build associations with their schools, teachers, and peers. As mentioned before, building relationships at school is a rehearsal and preparation to build connectedness with the real world. A well-prepared individual is more likely to adapt to a complex society [48]. However, experiencing bullying at school will make adolescents feel insecure, competitive, and even dangerous, so it will be very difficult for them to connect with school, and they will not feel supported, recognized, and valuable. As a result, it is not easy for them to positively adapt to the social environment.

Another important finding of the present study is that self-disclosure and school connectedness serve as serial mediators between school bullying and social adaptation during adolescence, which supports the fourth hypothesis. Experiencing school bullying will make adolescents less willing to disclose themselves. Sequentially, they might not build a connection with their school, which will influence their adaptation to society. To a certain degree, this finding provides support for the social penetration theory by stressing that self-disclosure is indeed conducive to building connections. Moreover, this finding extends prior studies by figuring out that, in addition to the independent mediating effects, self-disclosure and school connectedness jointly have a sequential mediating effect on the relationship between school bullying and social adaptation. Although the magnitude of the chain mediation is not the largest, it still has an efficient influence on the intervention to avoid the effects of school bullying for adolescents. Theoretically speaking, this finding encourages future studies to draw attention to further associations between mediators.

Similarly, a combination of the disclosure reciprocity effect and social penetration theory could also help better understand this finding [37, 53]. Knowing each other is a prerequisite for establishing a relationship. That is, self-disclosure plays a fundamental role in building connections with others. Self-disclosure will make adolescents feel supported, cared for, and encouraged at school. As a result, they will tightly connect with their peers, teachers, and schools. However, engagement in school bullying as bullies will make them feel the unequal power and status between themselves and others [1], which consequently will reduce the willingness of bullies to disclose themselves to others [31, 32]. Without self-disclosure, they will not build close connections with schools. More seriously, this kind of social alienation and withdrawal will have harmful impacts on their adaptation to society.

In summary, the present study makes several contributions to the research domain of school bullying. First, the present study extends the previous literature by directly investigating the influences of school bullying on adolescent positive and negative social adaptation. This direct finding repeatedly stresses the necessity to intervene in school bullying. Second, the present study figures out the three underlying mechanisms, which assist us in further understanding the relationship between school bullying and social adaptation. Practically, these mechanisms also provide more pathways for intervention in school bullying. Third, the sequential mediation effect in the present study suggests the interaction between the school subsystem and the individual subsystem in adolescence. This encourages future studies to focus on the complicated and dynamic associations between the environment and individuals.

Despite its unique advantages, the present study is not without limitations. First, with regard to participants, the present study has only recruited 434 Chinese junior high school students, which is not a large sample. Also, the findings cannot be generalized to other countries. So, future studies can include more participants from different cultural backgrounds. Second, all the data is collected by self-reported questionnaires, which cannot avoid the potential bias. Therefore, future studies can use other ways to collect data, such as combining other-reported questionnaires and other indicators. Third, the present study used a cross-sectional design, which cannot obtain causal results, but adolescent development is a dynamic process. Thus, future studies should use a longitudinal design to further explore the relationship between school bullying and social adaptation. Fourth, the present study did not involve any moderators to assess the stability of the relationship between school bullying and social adaptation in different situations. Compared to their counterparts with low self-control, adolescents with high self-control have fewer bullying behaviors and develop more positive outcomes, which indicates their high level of positive social adaptation to a certain degree [66, 67]. This suggests that future studies could attempt to evaluate the possible moderating role of self-control in the aforesaid relationship.

By constructing a serial mediation model, the present study demonstrated that school bullying has an influence on adolescent social adaptation directly as well as indirectly through the serial mediating mechanisms of self-disclosure and school connectedness. These findings highlight the importance of simultaneously examining self-disclosure and school connectedness as mediators between school bullying and adolescent social adaptation. Regarding interventions aimed to prevent or reduce adolescent school bullying, practitioners should pay more attention to the combined application of individual and environmental factors.

## Data Availability

All relevant data are within the manuscript and its Supporting Information files.

## Abbreviations

SB: School bullying;
PSA: Positive social adaptation;
NSA: Negative social adaptation;
SD: Self-disclosure;
SC: School connectedness.

## Declarations

## Ethics approval and consent to participate

This study was approved by the Research Ethics Committee of the College of Education and Sports Sciences, Yangtze University. All participants agreed to participate and provided informed consent.

## Consent for publication

Not applicable.

## Availability of data and materials

The datasets used and/or analysed during the current study are available from the corresponding author on reasonable request.

## Authors’ contributions

XG designed the work. R-JZ, HL, P-YW, XJ and C-SZ collected the data. R-JZ and G-XX analyzed the data. G-XX drafted and revised the manuscript.

## Acknowledgements

To the students, parents, teachers, and school staff who made this project possible.

